# VIGIL - Video In Geriatric Intervention Clinic: A randomised controlled feasibility trial protocol comparing face-to-face and video delivery of a specialist preoperative clinic for older people

**DOI:** 10.1101/2022.05.11.22274120

**Authors:** Emma Thorman, Andrea Joughin, Ben Carter, Philip Braude

## Abstract

The Covid-19 pandemic resulted in significant challenges to the provision of face-to-face clinics in geriatric perioperative care (G-POC). There are no studies evaluating the use of telemedicine in this population. A pilot study at North Bristol NHS Trust demonstrated that delivery of GPOC clinics via video consultation was feasible, but did not record outcome measures to demonstrate effectiveness and was not compared to face to face clinic. This study aims to provide proof of concept examining the outcomes of virtual G-POC consultations, compared to a face-to-face clinic, using standardised perioperative outcomes. It will test the feasibility of the intervention with a view to developing a randomised controlled trial.

## 1 Title of the study

VIGIL (Video In Geriatric Intervention cLinic): A randomised controlled parallel group feasibility trial protocol comparing face-to-face and video delivery of a specialist preoperative clinic for older people.

### Short Study Title

VIGIL - Video In Geriatric Intervention cLinic

## 2 Trial registration

IRAS Number: 310265

ISCRTN: 95833238

Sponsors Number: R&I 5110

Funders Number: Bristol Health Research Charity RF11

## 3 Protocol Version Number and Date

Version 1.4

Date 17^th^ February 2023

## 4 Signature Page

The undersigned confirm that the following protocol has been agreed and accepted and that the Chief Investigator agrees to conduct the study in compliance with the approved protocol and will adhere to the principles outlined in the Declaration of Helsinki, the Sponsor’s SOPs, and other regulatory requirement.

I agree to ensure that the confidential information contained in this document will not be used for any other purpose other than the evaluation or conduct of the investigation without the prior written consent of the Sponsor

I also confirm that I will make the findings of the study publicly available through publication or other dissemination tools without any unnecessary delay and that an honest accurate and transparent account of the study will be given; and that any discrepancies from the study as planned in this protocol will be explained.

**For and on behalf of the Study Sponsor:**

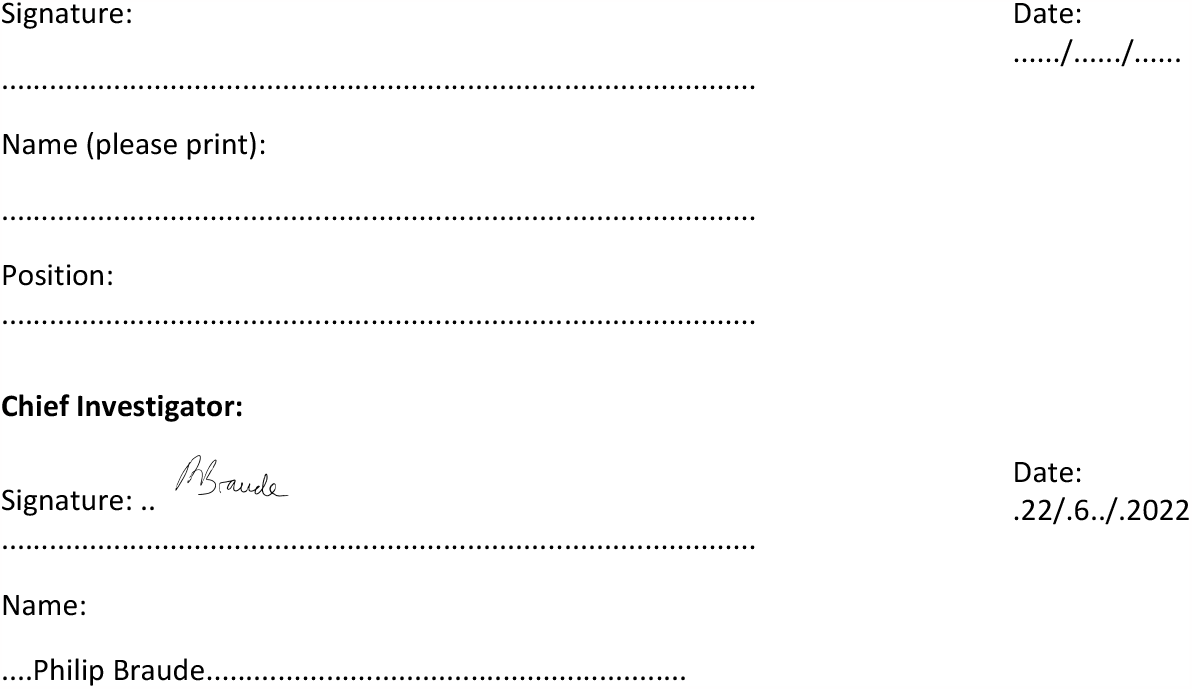

## 5 Key Study Contacts

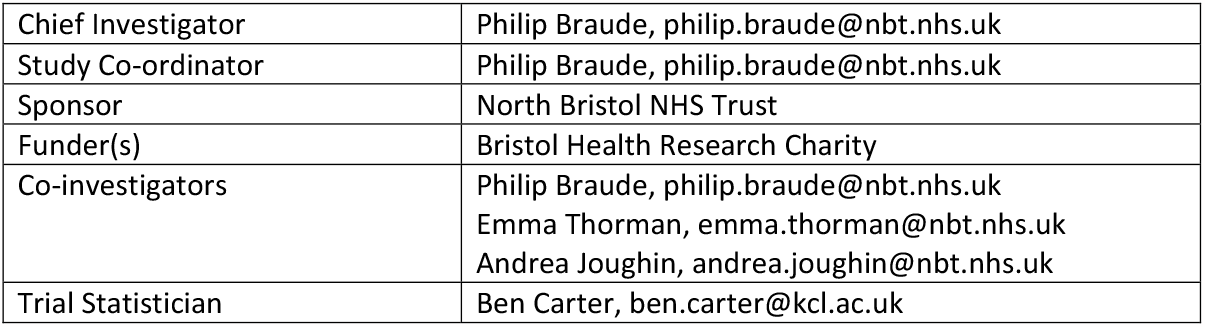

## 6 Study Summary

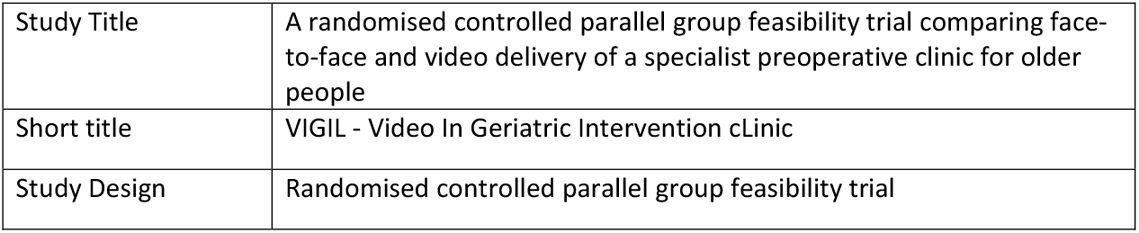
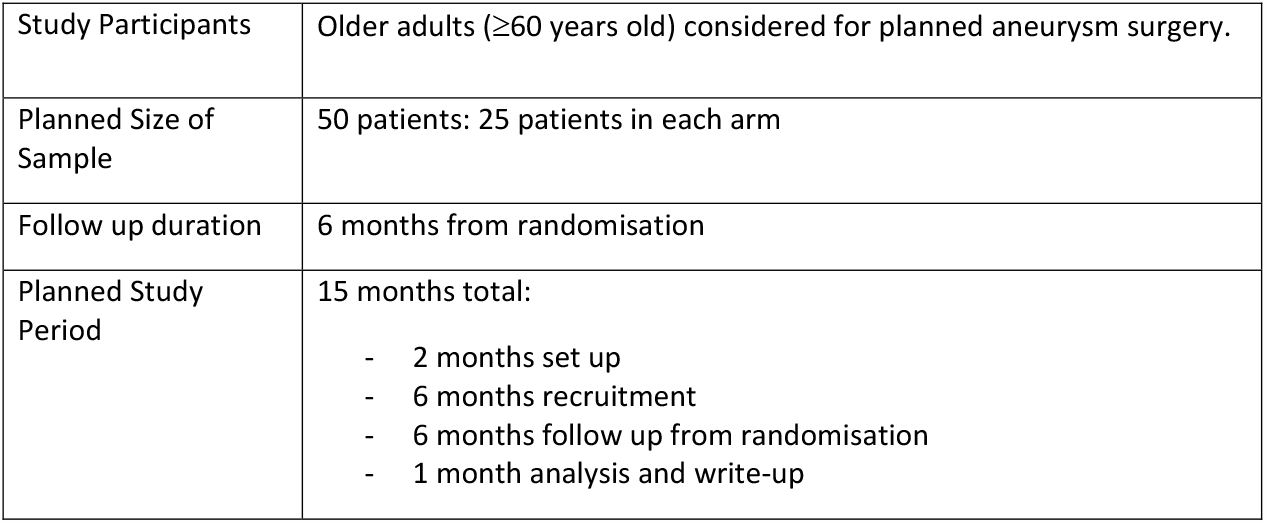

|  |  |
| --- | --- |
| Study Title | A randomised controlled parallel group feasibility trial comparing face-to-face and video delivery of a specialist preoperative clinic for older people |
| Short title | VIGIL - Video In Geriatric Intervention cLinic |
| Study Design | Randomised controlled parallel group feasibility trial |
| Study Participants | Older adults ($\geq 60$ years old) considered for planned aneurysm surgery. |
| Planned Size of Sample | 50 patients: 25 patients in each arm |
| Follow up duration | 6 months from randomisation |
| Planned Study Period | 15 months total:<br>- 2 months set up<br>- 6 months recruitment<br>- 6 months follow up from randomisation<br>- 1 month analysis and write-up |

### 6.1 Study Summary Outcomes

| Objective | Outcome |
| --- | --- |
| 1) To assess the feasibility of delivering a virtual geriatric preoperative clinic, compared to a face-to-face clinic. | - Number of patients randomised<br>- Proportion of patients adhering to the intervention allocated<br>- Proportion of patients followed up<br>- Proportion of patients with completed preoperative assessments including:<br>i. cardiac assessments including: blood pressure (in last 12 months), ECG (in last 12 months), exercise capacity (completion of Duke Activity Status Index), examination for heart failure (fluid balance assessment).<br>ii. cognitive assessments including: Telephone-Montreal Cognitive Assessment (MoCA)<br>iii. perioperative risk assessment including: Surgical Outcomes Risk Tool (SORT)<br>iv. respiratory assessments including: saturations (in last 12 months)<br>v. diabetes assessments including: HbA1c (within the last 3 months if patient has diabetes)<br>vi. Proportion of patients with additional or missed diagnoses compared to preoperative anaesthetic |
|  | assessment for example: cardiac - heart failure, valvular heart disease, ischaemic heart disease; respiratory – chronic obstructive pulmonary disease, interstitial lung disease. |
| 2) To assess the feasibility of measuring shared decision making | - Proportion of patients with completed shared decision making tool (collaboRATE)<br>- Proportion of patients that convert to surgery |
| 3) To test feasibility through process mapping of the preoperative pathway:<br>- Optimal time for recruitment<br>- Optimal time for delivery of the clinic | - Process map of the perioperative pathway<br>- Suggested time points for delivery of a video clinic |
| 4) To test the feasibility of collecting clinical outcomes of face-to-face and virtual clinics using standardized outcomes | - Proportion of patients with complete outcomes:<br>1) For all patients, surgery or not surgery<br>- mortality<br>- quality of life<br>2) For those that have surgery only<br>- length of hospital stay<br>- complications |

### 6.2 Plain Language Summary

The COVID-19 pandemic has forced many healthcare services to close. Millions of operations have been delayed globally. However, some urgent operations are still needed.

Older people having an operation at North Bristol Trust (NBT) are reviewed by a geriatrician (medical specialist in older people’s health). The review aims to improve health before surgery and support the patient’s decision making by exploring the risks and benefits of surgery, taking into account their underlying health and values. Few hospitals offer this service.

Due to the risk of hospital acquired COVID for this vulnerable patient group, we changed from a face-to-face clinic to a video clinic. The service received excellent feedback from patients and doctors in a study. However, we did not look at the effect on recovery after surgery or the feasbility

#### Study Aim

We intend to collect information before and after aneurysm surgery about complications and recovery. We would like to compare patients that had a geriatrician video appointment against face-to-face appointment. The point of this study is to find out if a large study could work to show if there is any difference between the two types of clinic. We want to get the process of running a trial right, and check whether patients are able and wish to take part. This smaller study will not show if video or face-to-face appointments are better than one another, but will help us to sort out the complicated process of running a large trial in the future.

#### Study Design

This is a randomised controlled trial meaning a computer system will decide which option of an appointment either via video call or face-to-face at Southmead hospital. The computer will select at random - this means there is an equal chance of getting either a face-to-face or video appointment.

#### Study Delivery

Patients referred from the vascular surgeons will receive a geriatrician face-to-face or video appointment as usual.

We will approach people to consent to the study who need planned aneurysm surgery. This will not include people due an urgent operation. It will also exclude those people who are unable to join a video clinic due to lack of equipment, their eyesight or hearing being too poor, or have severe communication difficulties.

The admin team will email or post study leaflets to patients interested. A doctor will call to answer questions and take consent. If a person lacks capacity (unable to make a decision) to consent to joining the trial, an appropriate carer or relative can provide advice to the study team if the person would have wanted to join. Consent will be checked at the appointment.

Information for the study will be collected before and after the person’s operation, while in hospital including any complications, and then after the operation to see if they survived the surgery.

An advisory patient group and stakeholder group will be setup to help design a future larger study.

#### How the information will be used

The results of this study will be offered to patients involved. The results will be published in a medical journal about older people and discussed at medical conferences.

We hope this study will pave the way to a larger study to show the benefits of video clinic. Video clinics can reduce the risk of hospital-acquired infection such as COVID-19, reduce travel with its associated financial and physical burden for patients as well as environmental impact, are delivered at times convenient for patients and doctors, and allow better access to special services for people across the UK.

## Data Availability

All data produced in the present study are available upon reasonable request to the authors

## 7 Funding and Support in Kind

| FUNDER(S) | SUPPORT GIVEN |
| --- | --- |
| Bristol Health Research Charity | Financial. Direct support for this study |
| Research Capability Funding from North Bristol Research and Innovation Department. | Financial. Not directly for this study. |
| NIHR Maudsley Biomedical Research Centre at the South London and Maudsley NHS Foundation Trust in partnership with King's College London | Financial. Not directly for this study. |

## 8 Role of Study Sponsor and Funder

Bristol Health Research Charity – supporting the development of this work through the Covid Research Fellowship. No direct influence over any part of the work.

North Bristol Trust R&I – supporting the development of this work. No direct influence over any part of the work.

## 9 Roles and Responsibilities

### 9.1 Study Steering Groups

|  | Role | Responsibility |
| --- | --- | --- |
| Trial management group (TMG) | Plan the day-to-day running of the trial. The TMG will consist of the CI and members of the research team including the site clinicians, the researchers, the trial manager, the trial statisticians, and the PPI. | Ensure the trial runs as per the protocol safely and identify any deviations. |
| Trial Steering group (TSC) | To provide study level oversight and review any safety events | Support the TMG with independent oversight |
| Stakeholder Committee | Members of the multidisciplinary team working within and interacting with the G-POC department at NBT. | Study development |

### 9.2 Protocol Contributors

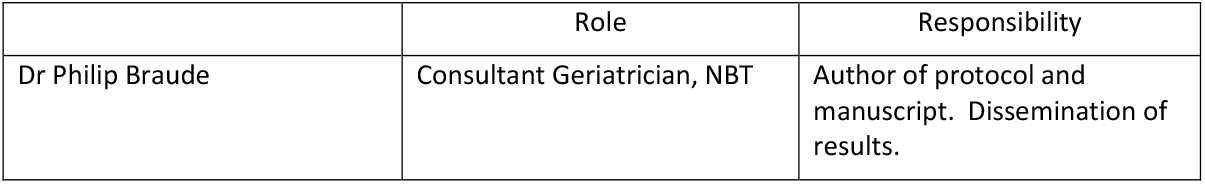
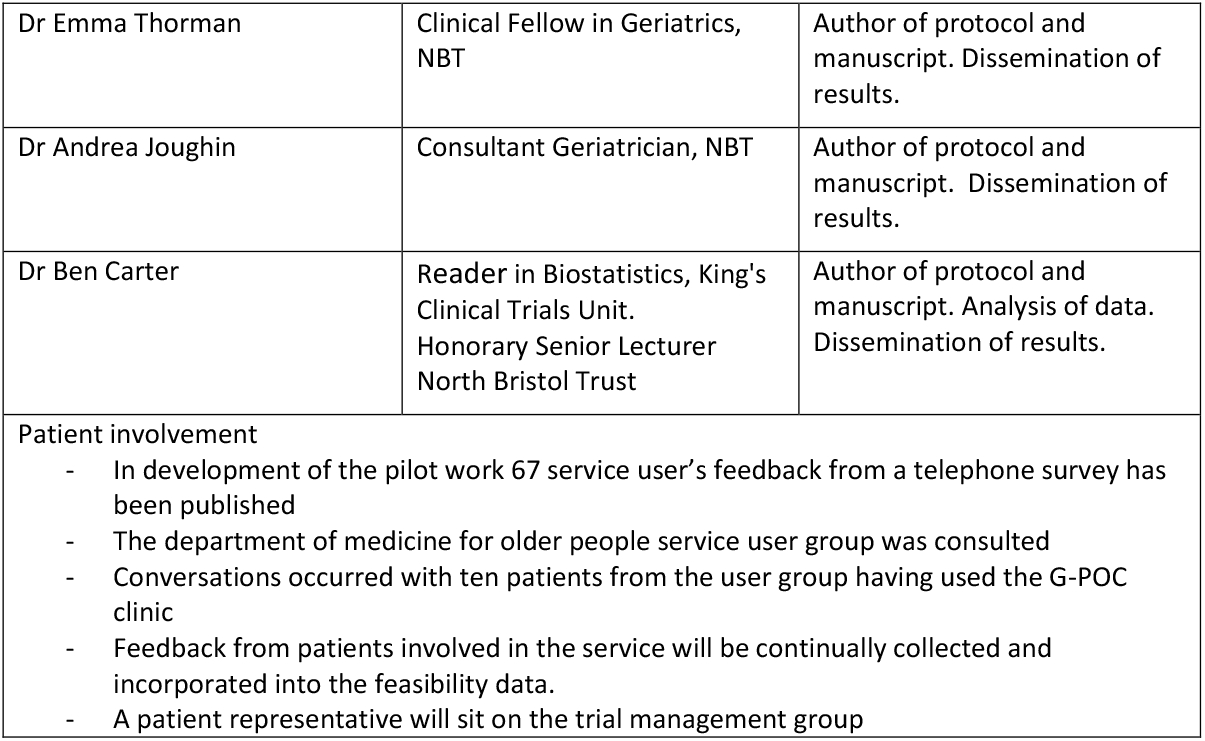

## 10 Study Flow Chart

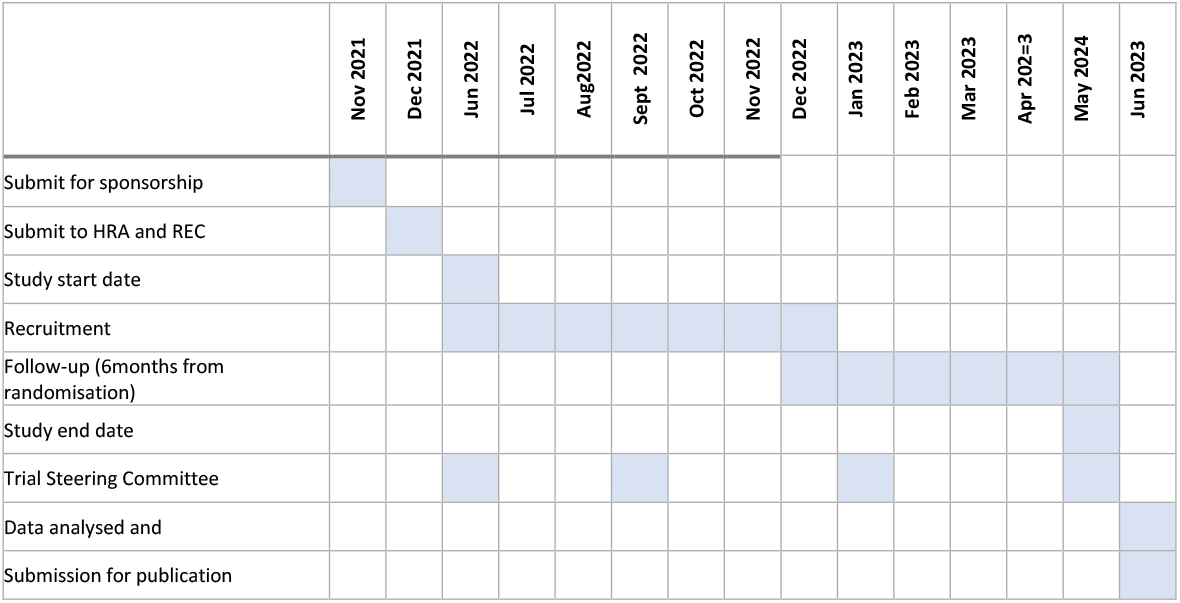

## 11 Background and Rationale

During the COVID-19 pandemic elective surgery has needed to be sacrificed for overwhelmed medical emergency services. An estimated 28,404,603 elective operations were postponed globally during the 12-week first peak (1). However, urgent elective surgical cases still needed to be prioritised with NHS England laying out guidance for those that should still be considered to proceed to surgery (2).

At NBT many of those referred for consideration of vascular surgery, and in particular aneurysm repair, were in a group to be prioritised. However, the usual preoperative pathway, incorporating a surgeon, anaesthetist, and geriatrician review, had to be altered due to a lack of usual face-to-face services and staff. In response, the geriatric perioperative care (G-POC) team piloted converting the face-to-face clinic to a new virtual video clinic. Just as with the face-to-face clinic, the aim of this clinic was to deliver optimisation of health and social circumstances prior to surgery, as well as to support patient-centred shared decision making.

The use of telemedicine has been evaluated prior to the pandemic and shown to be an effective tool with variable adoption. The VOCAL study (Virtual Online Consultations: Advantages and Limitations), pre-dating COVID-19, showed that virtual consultations can be safe and effective at delivering clinical services. They are popular with patients with less travel, and the ability to see patients in their own homes with relatives (3). Studies are underway to evaluate feedback on consultation process since COVID-19 in other specialties (4). Focusing on older people, a recent systematic review showed virtual clinics were effective for dealing with frailty issues such as polypharmacy and cognitive impairment (5). Only one of these studies compared outcomes of a virtual to face-to-face consultation demonstrating virtual clinics had a greater capacity for seeing patients more rapidly, and therefore delivery on a greater scale, with similar reductions in health service usage compared to a face-to-face visiting geriatrician review (6).

No literature exists in perioperative medicine for older people around video clinics. With this evidence gap we undertook a quality improvement evaluation. A new virtual clinic assessment was tailored using Plan-Do-Study-Act cycles. 67 patients being considered for vascular (59 patients) or cancer surgery (8 patients) in COVID Wave 1 were reviewed virtually: video consultation 43%, telephone 57%. Barriers, facilitators, and patient feedback were evaluated. The study, published in Age and Ageing May 2021 (7), reported that almost all participants (90.6%) reported understanding the reason for the consultation, and felt better able to manage and understand their condition. Clinicians rated virtual consultations as able to deliver preoperative assessment, medical optimisation, and shared decision-making (video consultations 89.7%, telephone consultation 68.4%).

Despite our pilot work setting up the virtual clinic and demonstrating feasibility in delivery, we did not record any outcome measures to demonstrate effectiveness. In addition, no face-to-face clinic was available to be evaluated alongside.

The establishing proof of concept for G-POC outpatient video consultations will inform the development of the study protocol for a larger randomised controlled trial.

This study aims to provide proof of concept examining the outcomes of a standard-of-care preoperative virtual geriatric clinic, compared to a face-to-face clinic, using standardised perioperative outcomes. It will test feasibility of the intervention with a view to developing a fully powered randomised controlled trial

## 12 Objectives

### 12.1 Primary objective

1. To assess the feasibility of delivering a virtual geriatric preoperative clinic, compared to a face-to-face clinic.
  a. To test feasibility of randomising
  b. To test adherence to the allocated groups
  c. To test the feasibility of follow-up
  d. To test the feasibility of delivering a standardised preoperative assessment

### 12.2 Secondary objectives

1. To assess the feasibility of measuring shared decision making
2. To test feasibility through process mapping of the preoperative pathway:
  a. To explore the optimal time for recruitment
  b. To explore the optimal time for delivery of the clinic
3. To test the feasibility of collecting clinical outcomes of face-to-face and virtual clinics using standardized outcomes:
  c. For all patients, surgery or not surgery: quality of life, mortality.
  d. For those that have surgery only: length of hospital stay, complications.

This may also help in identifying any early signals between the two groups with a view to sample size calculations and developing primary outcomes to focus on in a future fully powered randomised controlled trial.

## 13 Trial Design

Randomised controlled trial with two groups compared:

- Interventional group: Standard of care video preoperative clinic.
- Control group: Standard of care face-to-face preoperative clinic.

### 13.1 Study Setting

This is a single centre study based at North Bristol NHS Trust (NBT), a tertiary referral hospital and major trauma centre in the South West of England. It is also the home of the Major Arterial Centre, which brings together all inpatient elective and emergency arterial surgery for the Bristol Bath Weston Vascular Network. The geriatric perioperative care (GPOC) clinic delivers pre-operative assessment, personalised risk assessment, medical optimisation and shared decision making for older people considering surgery.

### 13.2 Eligibility Criteria

#### 13.2.1 Inclusion criteria

- proposed aneurysm surgery
- be able to read and communicate in English
- 60 years old or over

#### 13.2.2 Exclusion criteria

- emergency surgery prior to clinic attendance (within 1 week of referral)
- inability to participate in video consultation due to:
  - lack of access to appropriate technology
  - significant sensory impairment: registered blind or functional severe sight impairment e.g. unable to read study materials, or auditory e.g. unable to communicate using the telephone on screening
  - inability to communicate on the telephone at screening e.g. advanced dementia

### 13.3 Interventions

Following consent patients will be randomised into one of the two groups:

1. Face to face G-POC appointment (control)
2. Virtual G-POC appointment (intervention)

Patients will receive a standardised comprehensive geriatric assessment (CGA). There will be natural variation in the manner of conducting a CGA for each clinician, however, this will be standardised by:

1. Using domains following the Frailty Domains as proposed by Centre for Perioperative Care Guideline for Perioperative Care for People Living with Frailty Undergoing Elective and Emergency Surgery (8)
2. Using standardised assessment tools for each domain
3. Employing appropriate guidance to each domain

In order to ensure safety in the intervention arm, an additional face-to-face backup appointment will be arranged in case the virtual appointment is inadequate to complete the assessment. This additional appointment will be on a day the patient is already attending Southmead hospital. In practice this will work with patients being booked in for four appointments back-to-back on a Wednesday morning: vascular surgery, nurse-lead preoperative assessment, cardiopulmonary exercise testing, and G-POC face-to-face clinic. The intervention group will have a virtual appointment prior to this half day of face-to-face appointments. If the virtual appointment is completed successfully then the patient will not need to attend the G-POC face-to-face appointment and it will be cancelled.

### 13.4 Outcomes

#### 13.4.1 Primary outcome

1. To assess the feasibility of delivering a virtual geriatric preoperative clinic, compared to a face-to-face clinic.
  a. Number of patients randomised: The number of patients willing to be randomised to different modalities of delivering the clinic. Success is defined as 50 patients, partial success if 80% are randomised (40 or more patients).
  b. Proportion of patients adhering to the intervention allocated
  c. Proportion of patients followed up
  d. Proportion of completed preoperative assessments:
    i. cardiac assessments including:
      1. blood pressure (in last 12 months) (9)
      2. ECG (in last 12 months) (10)
      3. exercise capacity (completion of Duke Activity Status Index) (11)
      4. examination for heart failure (fluid balance assessment)
    ii. cognitive assessments including: Telephone-Montreal Cognitive Assessment (T-MoCA) (8)
    iii. perioperative risk assessment including: Surgical Outcomes Risk Tool (SORT)
    iv. respiratory assessments including: saturations (in last 12 months)
    v. diabetes assessments including: HbA1c (within the last 3 months if patient has diabetes) (10)
    vi. Proportion of patients with additional or missed diagnoses compared to preoperative anaesthetic assessment for example: cardiac - heart failure, valvular heart disease, ischaemic heart disease; respiratory – chronic obstructive pulmonary disease, interstitial lung disease.

#### 13.4.2 Secondary outcomes

2) To assess the feasibility of measuring shared decision making
  a. Proportion of completed shared decision making tool (collaboRATE) (12)
  b. Proportion of patients that convert to surgery
3) To test feasibility through process mapping of the preoperative pathway:
  a. Process map of the perioperative pathway to determine optimal time for recruitment
  b. Suggested time points for delivery of a video clinic
4) To test the feasibility of collecting clinical outcomes of
  - For all patients surgery or not surgery
    - mortality (at discharge and 6 months post-randomisation)
    - quality of life (at 6 months post-randomisation)
  - For those that have surgery only
    - length of hospital stay
    - complications (Postoperative Morbidity Score at inpatient days 1, 3, 5, 8)

### 13.5 Sample Size

Consistent with feasibility studies 50 patients will provide adequate data to estimate the key parameters needed for a fully powered RCT. 25 patients will be recruited into each arm of the trial.

## 14 Recruitment

See recruitment and consent flow chart page 15.

The trial will aim to recruit patients awaiting aneurysm surgery who are referred to the G-POC clinic during the study period. At the point of referral to the clinic, during routine booking for G-POC clinic patients will be screened against the eligibility criteria by G-POC administrators or a member of the study team.

### 14.1 Screening

Patients will be referred by the usual clinic pathway to the G-POC clinic by their surgeon, anaesthetist, or as part of the agreed standard pathway of care for vascular surgical patients.

Screening will use a standardised form which will capture:

- Proposed surgery
- The ability of the participant to engage in a video clinic

Those meeting the inclusion criteria will be:

- invited to take part in the study in an initial consent telephone call on the same day completed by a member of the study team
- sent via email or by first class post:
  - i) a patient information leaflet ii) a blank consent form
- contacted by a member of the study team two days later by telephone to:
  - answer any questions
  - provide information about the trial
  - take consent to participate in the study
  - complete a Clinical Frailty Scale to be used for randomisation

This process will minimise delays in booking the patient into their G-POC clinic. It is crucial to minimise delays within the clinical pathway as patients will be on treatment pathways with hospital treatment targets, and often with pathology requiring urgent or expedited decisions for surgery, for example a large aneurysm at risk of rupture (enlarged major blood vessel at risk of bursting).

### 14.2 Consent

#### 14.2.1 Consent for patents who have capacity

##### At recruitment

The default method will be to take consent over the telephone. It will not be possible to take initial consent in person due to the nature of the study requiring randomisation before the patient is seen face-to-face or by video.

The clinician will initial the consent form in relevant sections to indicate each area has been discussed and agreed to by the patient. The consent form will be kept in the site file, a copy posted to all study participants for their records, and sent to their GP.

##### At study follow-up

The participant’s agreement to continue participation in the trial will be checked at each stage of study:

- rechecked at the beginning of the video or face-to-face appointment
- rechecked face-to-face at the start of inpatient data collection
- rechecked by telephone at the start of outpatient follow-up

#### 14.2.2 Consent for patients who lack capacity

Patients who lack capacity are less likely to proceed to surgery in this vascular cohort of patients. Comorbid disease such as dementia is not uncommon. Such patients have limited gains from aneurysm surgery in the context of a life limiting neurodegenerative condition. However, patients may still benefit from the preoperative assessment process and shared decision making.

If patient is unable to understand, retain, weigh up, or communicate a decision to be involved in the study, and therefore lack capacity, a personal consultee will be sought to garner advice on whether they feel their relative/friend would want to participate in the trial. The personal consultee will be provided with a Consultee Information Sheet. Questions will be answered over the telephone. If they are happy to take on the role a Consultee Declaration Form will be complete on the telephone. They will then be asked if they would advise if they feel their relative/carer would wish to participate in the trial. If they are not happy to take on the role then another personal consultee will be sought. If no personal consultee can be found then the person will not be recruited into the trial.

If the person regains capacity then they will be approached to take consent in the default method.

#### 14.2.3 Patients who lose capacity

Patients may lose capacity to agree to continue their involvement in the study. If this occurs it is most likely to happen after surgery secondary to postoperative delirium. About 20-30% of vascular patients will have a postoperative delirium. This does not automatically mean the participant will lack capacity if they have delirium.

A statement as to the possibility of losing capacity to agree to the study follow-up will be included in the Patient Information Sheet and Consent Form. This will provide support for continuing follow-up even if capacity is lost to continue in the study. In addition contact details for a personal consultee will be sought at the consent process. If the participant loses capacity the personal consultee will be contacted to ensure they are in agreement that the participant may continue in the study.

### 14.3 Randomisation

The allocation sequence will be generated using a varying permuted block design stratified by frailty status using the clinical frailty scale (CFS 1-4, 5-6, 7-8). The sequence will be masked from recruiting members of the study team prior to and on the day of the baseline assessment.

The enrolling clinician will conduct the baseline assessment with a frailty score and will enter the study data onto the study database. They will then inform the study statistician who will allocate the patient on the following day.

Randomisation will take place using a sealed envelope system setup by the study methodologist. At the point of randomisation the blinded study methodologist will be contacted to provide the group allocation.

### 14.4 Blinding of participants

Patients will not be blinded to the intervention due to the nature of the intervention. The study team will be unblinded to the allocation. No mechanism is required for emergency unblinding as the Chief investigator will be unblinded.

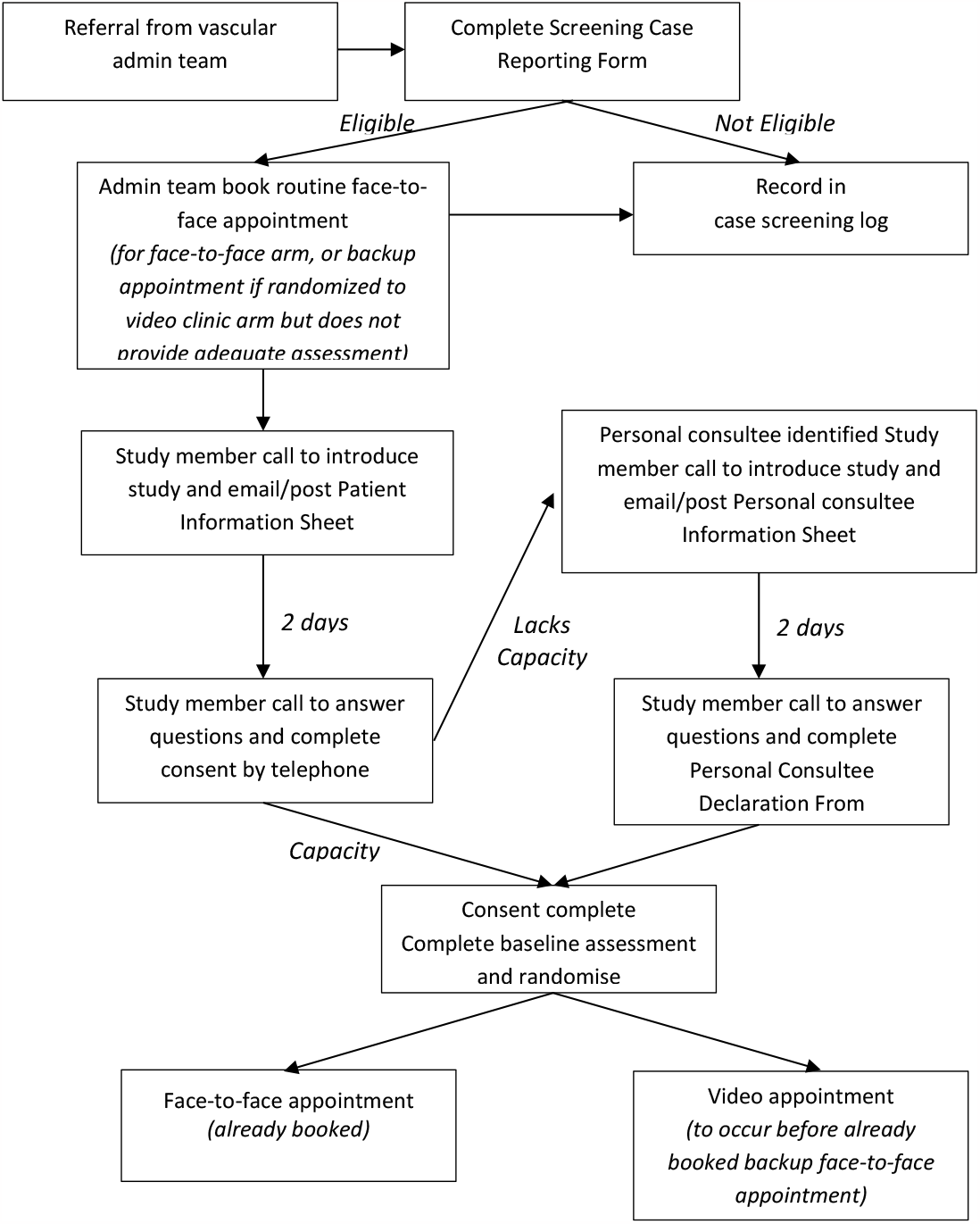

## 15 Methods

### 15.1 Data collection methods

Data will be collected

- screening by referral information and telephone assessment
- baseline by either video or face-to-face assessment as randomised
- inpatient by face-to-face assessment
- 2 days (+7 days) post intervention via telephone call
- Follow up six months post-randomisation (+/-2 weeks) by telephone.

### 15.2 Sampling technique

Study participants will be identified using convenience sampling of patients awaiting major vascular surgery referred to G-POC clinic.

### 15.3 Trial data collection

Data will be collected at 5 time points using a standardised booklet to complete the clinical information. These booklets are used in the clinic already for clinicians to document their findings. A standardised case reporting form will be used for additional data that are additional to usual standard of care (denoted in the table below).

### 15.4 Timing of Assessments

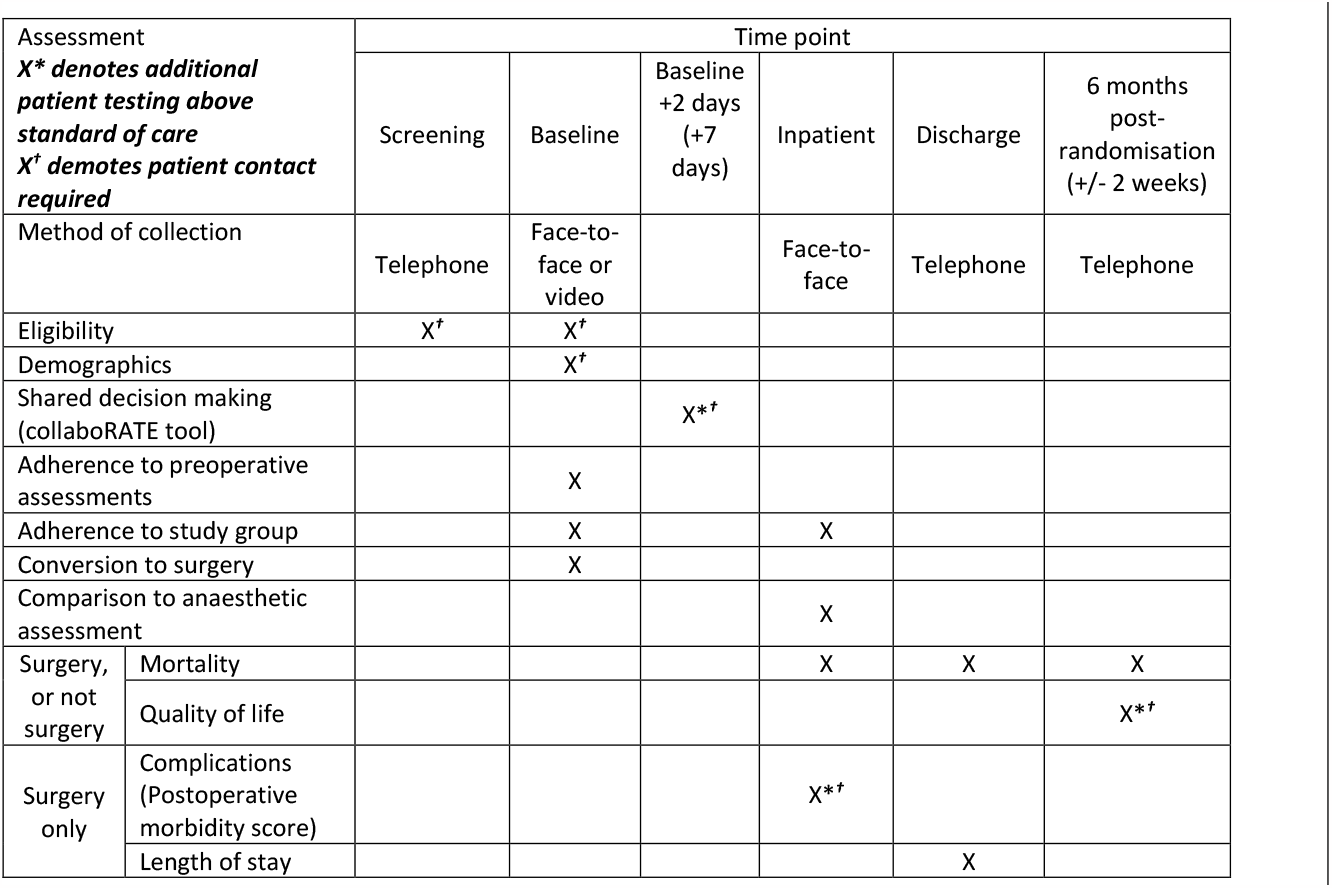

### 15.5 Description of Assessments

Demographics will include: patient age, sex, ethnicity, index of multiple deprivation (using postcode in the UK Government English indices of deprivation 2019 website).

Standard of care assessments will include outcomes detailed in the trial requiring no additional data collection: cardiac assessment, respiratory assessment, diabetes assessment, cognitive assessment, risk assessment.

Shared decision making will be determined using the collaboRATE 5 point scale tool. This will be completed with the patient by telephone with a member of the study team that was not involved in the clinical assessment. This will minimise bias of the patient completing an evaluation of the consultation with the clinician who delivered the consultation. If a personal consultee has been nominated the “collaboRATE for individuals acting on behalf of patients” will be considered to be used.

Adherence will be determined by patients that receive the intervention as allocated without additional data collected outside of the clinic appointment.

Comorbidity will be collected using the Charlson Comorbidity Index.

Postoperative assessments will include: length of hospital stay, complications using the Postoperative Morbidity Score (13).

Adverse Events will be asked at each time-point using an unstructured log.

### 15.6 Process mapping

Visual diagrams of the steps of the preoperative pathway will be created using computer software. This will include stages of:

1. referral into the preoperative pathway e.g. GP referral, aneurysm screening program
2. referral into the G-POC clinics
3. review by other clinicians e.g. surgeon, anaesthetist, preoperative nurse
4. pathway to surgery.

## 16 Data management

A unique identifier will be assigned to each participant on inclusion in the study. Source data will be the collected using standardised case report forms. Data will be entered onto a study database held on a secure server within the North Bristol Trust systems. Computers will only be able to be accessed using password protected user details. The electronic site files will only be accessible by those within the study team on a password encrypted database. At the end of the trial the Chief Investigator will approve the study data as complete and the data will be locked.

Data extracts will be provided to the statistician throughout the trial who will store the data anonymously on a secure server. The statistician will code all instruments following the validated instrument scoring methods.

A physical and electronic site file will be kept with all study materials, completed consent forms and case reporting forms. Any part of the physical site file will be held in a locked filing cabinet on the Southmead premises which is key card access only, the electronic site file will be held on the North Bristol Trust servers.

Data will be kept for three years after the study’s publication in order to answer any queries. After this date the data will be destroyed.

## 17 Statistical methods

### 17.1 CONSORT Flowchart

The CONSORT flow chart will summarise the population screened, those excluded and those randomised. Reasons will be provided to exclusion. Of those randomised all eligible patients will be followed up at each timepoint. Postoperative assessments will only be carried out on patients who have an operation.

### 17.1 Primary Feasibility Outcomes

The feasibility outcomes will be analysed using descriptive methods, eg as a frequency, or proportion. Proportions will be summarised with a 95% confidence interval. No hypothesis testing or p-values will be carried out on the feasibility outcomes.

### 17.2 Secondary Clinical Outcomes

Secondary outcomes of the clinical measures will be summarised with a between allocated group adjusted mean difference (aMD) for all continuous measures using a mixed effects multivariable regression, with a random effect fitted for patient and adjusted for the following fixed effects: baseline disease severity, patient age group, sex and CFS. The aMD will be fitted alongside the 95% confidence interval (95%CI), but no p-value will be presented.

For clinical categorical measures an adjusted odds ratio (aOR) will be presented with the 95% CI in a similar manner using a mixed effects multivariable logistic regression model.

#### Secondary Analysis of the secondary clinical outcomes

In addition to the secondary analysis of the clinical outcomes they will be also adjusted by: comorbidity, baseline disease severity.

#### Subgroup analyses

A subgroup analysis will be carried out on the only those patients that receive surgery.

#### Analysis Populations

The primary population under investigation will be the modified intention to treat population (mITT). The mITT population will include all randomised and eligible patients who complete the surgical assessment. Patients who do not have surgery will be matched by age to those who have surgery. This non-surgical group will be followed-up at the same time point. Missing data will be explored, but will not be imputed.

#### Interim analysis and stopping rules

No interim analysis is planned.

## 18 Harms

Adverse events (AEs) will be assessed using an unstructured AE log and asked at each visit. Whilst not expected, any serious AEs will be presented to the Trial Study Committee chair if the Chief Investigator records any SAE is related to the allocation arm. Harms will be reported to the study sponsor and documented in line with Good Clinical Practice.

Specific adverse events will include: cancelled surgery, postponed surgery

Specific serious adverse events will include: hospitalised emergency procedure, life threatening condition, death,

## 19 Trial Management

### 19.1 Study Sponsor

The protocol will have been reviewed and sponsored by the study site – North Bristol Trust.

### 19.2 Trial Steering Committee

The TSC will provide independent oversight of the study. Will meet at the beginning and end of the study to ensure the protocol is being and has been adhered to throughout.

### 19.3 Patient involvement group

The patient involvement group was identified through the G-POC service user group. Members have already flagged their availability to participate in research studies. Members of the group will not be recruited to the study. The group consisted of two members. Feedback and design of the protocol was also collected from a range of conversations with the G-POC service user group.

Patient involvement group will be asked to input at four time points:

1. Study protocol and documentation including assessment tool acceptability, outcome measure acceptability
2. Interpretation of the data
3. Writing of the manuscript and dissemination
4. Feedback at the end of the trial

### 19.4 Stakeholder group

The stakeholder group was identified as key participants involved in the perioperative pathway of vascular patients. Participants in the stakeholder group are independent of the study team. They included a vascular surgical consultant, anaesthetic consultant, cardiology consultant, G-POC patient administrator, vascular surgical administrator.

The stakeholder group will be asked to input at four time points:

1. Study protocol and documentation including assessment tools used, outcome measure examined, willingness to deliver randomised clinic care.
2. Interpretation of the data
3. Writing of the manuscript and dissemination
4. Feedback at the end of the trial

## 20 Ethics and dissemination

### 20.1 Research ethics approval

HRA and REC approval will be sought prior to the initiation of the study.

The protocol has been reviewed by the Chief Investigator and study co-ordinators, as well as the PPI group, prior to be being sent for assessment by the nominated Clinical Trials Officer in the Research Office at NBT.

### 20.2 Dissemination

The completed study will be submitted for peer review journal publication, social media including Twitter, scientific blog posts, national media including radio, television, and online. It will be presented at national conferences and submitted for oral and written abstracts. A study report will be written for the Bristol Health Research Charity funders.

## 21 Protocol Amendments

Accidental protocol deviations can happen at any time. They must be adequately documented on the relevant forms and reported to the Chief Investigator and Sponsor immediately.

Deviations from the protocol which are found to frequently recur are not acceptable, will require immediate action and could potentially be classified as a serious breach.

## 22 Confidentiality

All investigators and study site staff must comply with the requirements of the Data Protection Act 1998 with regards to the collection, storage, processing and disclosure of personal information and will uphold the Act’s core principles.

## 23 Declaration of interests

There are no competing interests.

## Notes

### Competing Interest Statement

The authors have declared no competing interest.

### Clinical Trial

ISRCTN95833238

### Funding Statement

This study has received funding from the Bristol Health Research Charity

### Author Declarations

Research Ethics Committee of the Health Research Authority and Health and Care Research Wales gave ethical approval for this work.

### Summary of Updates

Author list updated.

